# The value of Gene Xpert MTB/RIF of bronchial lavage fluid in the diagnosis of sputum AFB negative pulmonary tuberculosis

**DOI:** 10.1101/2025.04.06.25325327

**Authors:** Ngo The Hoang, Nguyen Duy Cuong, Pham Minh Tri, Phung Thao My, Do Thanh Son, Le Thi Diep, Dung Si Ho, Quoc Bui, Le Dinh Thanh, Thach Nguyen, Minh Huu Nhat Le

**Author notes:** ***Emails***: Ngo The Hoang, Nguyen Duy Cuong, Pham Minh Tri, Phung Thao My, Do Thanh Son, Le Thi Diep, Dung Si Ho, Le Dinh Thanh, Thach Nguyen., Minh Huu Nhat Le. or. Correspondent Authors*: ***Dr. Ngo The Hoang, MD***., – Affiliation: Department of Respiratory, Thong Nhat Hospital, Ho Chi Minh City 70000, Vietnam; or ***Dr. Minh Huu Nhat Le, MD, PhD***, or; *Affiliation*: International Ph.D. Program in Medicine, College of Medicine, Taipei Medical University, Taipei 110, Taiwan; Cardiovascular Research Laboratories, Methodist Hospital, Merrillville, IN; ***Dr. Thach Nguyen, MD***., *Affiliation*: Cardiovascular Research Laboratories, Methodist Hospital, Merrillville, IN 46410, United States.

## Abstract

**Objective:** To describe the characteristics of pulmonary tuberculosis patients with acid-fast bacilli (AFB) negative sputum and determine the value of Gene Xpert MTB/RIF of bronchial lavage fluid in diagnosis of sputum AFB-negative pulmonary tuberculosis.

**Method:** This cross-sectional study was conducted on 120 pulmonary tuberculosis-suspected patients with AFB-negative sputum admitted to the Respiratory Department of Thong Nhat Hospital from July 2022 to August 2023.

**Results:** A total of 56 patients were recruited, 67.9% were male and 32.1% female. Mean age was 57.8 ± 20.1 years and mean body mass index was 21.4 ± 1.8 kg/m^2^. Common comorbidities included hypertension (41.1%), type 2 diabetes (30.4%), and chronic lung disease (5.4%). 48.2% of patients had more than 1 lesion on X-ray. Lesion locations detected in X-ray were in right lung (46.4%), left lung (28.6%), upper lobe (51.8%), lower lobe (17.8%). Infiltrative injury on X-ray accounted for 64.3%, followed by consolidation (21.4%) pleural effusion (10.7%), cavity (8.9%), and nodular (5.4%). Gene Xpert MTB/RIF of bronchial lavage fluid found tuberculosis bacteria in 40.8% of cases. The positive rate of Mycobacteria growth indicator tube culture (MGIT) of bronchial lavage fluid was 46.7%. The sensitivity of Gene Xpert MTB/RIF compared to MGIT was 85.7% and the specificity was 98.4%. The positive predictive value was 97.9% and the negative predictive value was 88.7%. Rifampicin resistance rate was 4.1%.

**Conclusion:** Pulmonary tuberculosis patients with AFB-negative sputum have diverse clinical and radiological characteristics. Gene Xpert MTB/RIF of bronchial lavage fluid had high sensitivity and specificity in diagnosis of AFB-negative pulmonary tuberculosis. Rifampicin resistance rate was low.

## 1. INTRODUCTION

Tuberculosis (TB) is an infectious disease caused by *Mycobacterium tuberculosis*. Tuberculosis can be found in all parts of the body, of which pulmonary tuberculosis is the most common form (accounting for 80-85% of total cases) and is the main source of infection to surrounding people through droplets from the air. There were 10.8 million people infected with TB worldwide in 2024, equal to 134 new cases per 100,000 people ^1^. Around 400 thousand new cases of drug-resistant TB developed in 2023, continuing a stable trend since 2020 ^1^. This figure included rifampicin-resistant and multidrug-resistant TB, the latter also resistant to isoniazid ^1^. In Vietnam, tuberculosis incidence was estimated at around 100 cases per 100,000 adults in 2005-2015, and prevalence at 322 cases per 100,000 adults ^2,3^. Pulmonary TB prevalence was high at around 79 cases per 100,000 adults ^3^. Diagnosis of pulmonary TB in Vietnam is reliant upon clinical and chest X ray characteristics, as well as AFB sputum smear, with limited availability of sputum culture test for AFB smear negative but suspected cases ^4^. The efficacy of AFB sputum smear method in detecting TB might be as low as 50%, and dependent on local equipment quality ^5^. These conditions highlight the need for more effective diagnostic tools for TB in Vietnam.

The Gene Xpert MTB/RIF testing system was recommended by WHO for use in the initial diagnosis of suspected multidrug-resistant or HIV-associated TB in 2010, then expanded to all suspected TB cases in 2014 ^6^. The sensitivity of Gene Xpert MTB/RIF in sputum-negative TB was up to 67%, positioning it as a solid option to strengthen TB detection in Vietnam ^6^. To increase the accuracy in diagnosing pulmonary TB with AFB-negative sputum, we conducted this study with the following goals: (1) Describe the characteristics of pulmonary tuberculosis patients with AFB-negative sputum and (2) Determine the value of Gene Xpert MTB/RIF test in diagnosis of AFB-negative pulmonary tuberculosis at Thong Nhat hospital.

## 2. METHODS

### 2.1. Study design and participants

A cross-sectional study was conducted at Department of Respiratory in Thong Nhat Hospital, Ho Chi Minh City, Vietnam. There were 120 participants enrolled in the study from July 2022 to August 2023.

The inclusion criteria included (1) adult patients (≥18 years old) with clinical symptoms and/or lesions suggestive of tuberculosis on X-ray and (2) ≥2 AFB-negative sputum samples taken on the first day of admission (AFB sputum test was performed at the Microbiology Department of Thong Nhat Hospital).

The exclusion criteria were (1) severely ill patients, or (2) not agreeing to participate in the study, or (3) not providing qualified sputum samples.

### 2.2. Procedures

#### 2.2.1. Data collection and clinical examination

Participants provided personal information, with particular emphasis on medical history, including age, gender, smoking and alcohol consumption habits, history of tuberculosis, and other comorbidities. A comprehensive clinical examination was conducted for all participants.

#### 2.2.2. Laboratory and imaging tests

Diagnostic assessments included chest X-ray imaging, complete blood count analysis, liver and kidney function tests, prothrombin time, and activated partial thromboplastin time evaluations.

#### 2.2.3. Bronchial lavage sample collection

Flexible bronchoscopy was performed to collect bronchial lavage fluid samples for Mycobacteria Growth Indicator Tube (MGIT) culture and Gene Xpert MTB/RIF testing.

Absolute contraindications for flexible bronchoscopy included acute respiratory failure (unless the patient was intubated and mechanically ventilated), severe tracheal obstruction, inability to maintain adequate oxygenation during the procedure, and untreated or life-threatening arrhythmias.

Relative contraindications included recent myocardial infarction, uncooperative patients, and uncorrected coagulation disorders.

#### 2.2.4. Gene Xpert testing

Gene Xpert MTB/RIF testing was conducted using the Gene Xpert Cepheid machine (USA) equipped with the Xpert MTB/RIF GX IV-R2 cartridge model. The assay provided rapid results within two hours, determining the presence of tuberculosis bacteria and detecting rifampicin resistance.

### 2.3. Statistical analysis

Data was processed with SPSS 20.0 for Windows software. Continuous variables were presented as mean ± standard deviation (SD) for normally distributed data. Categorical variables were presented as frequencies (%). Chi-square or Fisher’s exact test was used to compare proportions, T-test for continuous variables with normal distribution. Statistical significance was accepted at p-value <0.05.

### 2.4. Ethics issues

The study was approved by the Board of Directors and the Ethics Committee of Thong Nhat Hospital (Reference No. 36/2022/CN-BVTN-HDDD) on April 26, 2022. Research period: July 2022 to August 2023. The requirement for informed consent was waived due to the retrospective, cross-sectional design. Patient data privacy and confidentiality were strictly maintained. The study supported timely and guideline-based diagnosis and treatment of patients with suspected AFB-negative tuberculosis.

## 3. RESULTS

During the study period, there were 120 patients with suspected pulmonary tuberculosis but had AFB-negative sputum. Only 56 patients had MGIT of bronchoalveolar lavage detecting tuberculosis with a mean age was 57.8 ± 20.1 and the body mass index (BMI) was 21.4 ± 1.8 kg/m2 (Table 1).

**Table 1.**
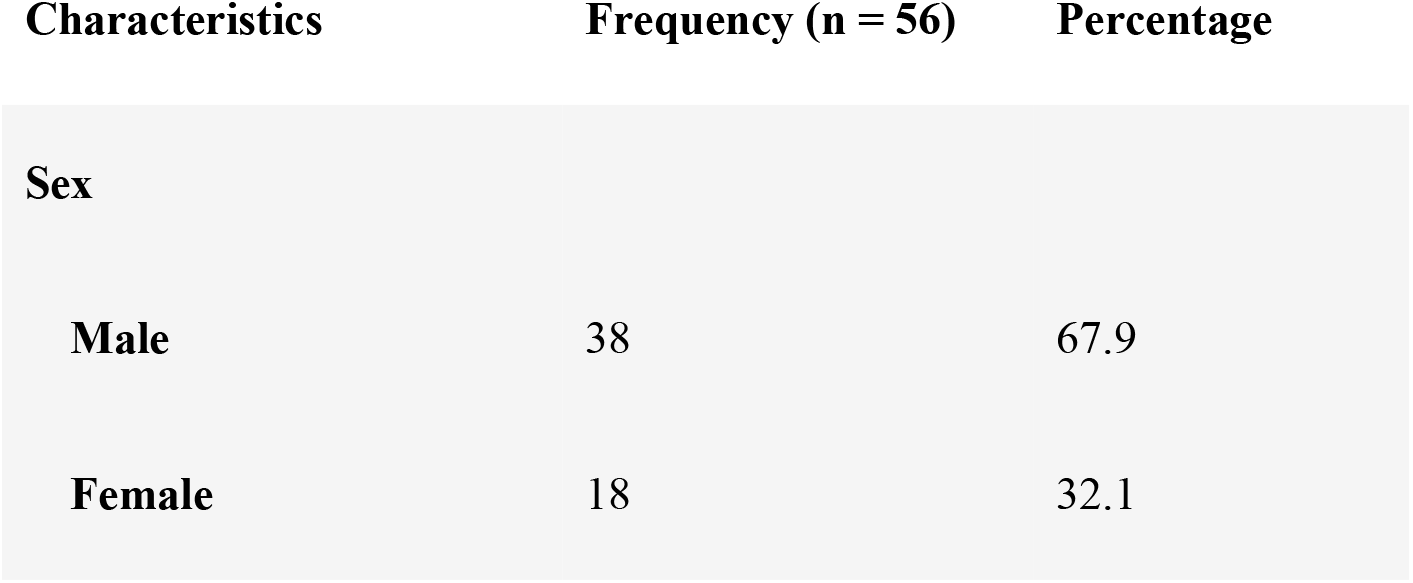
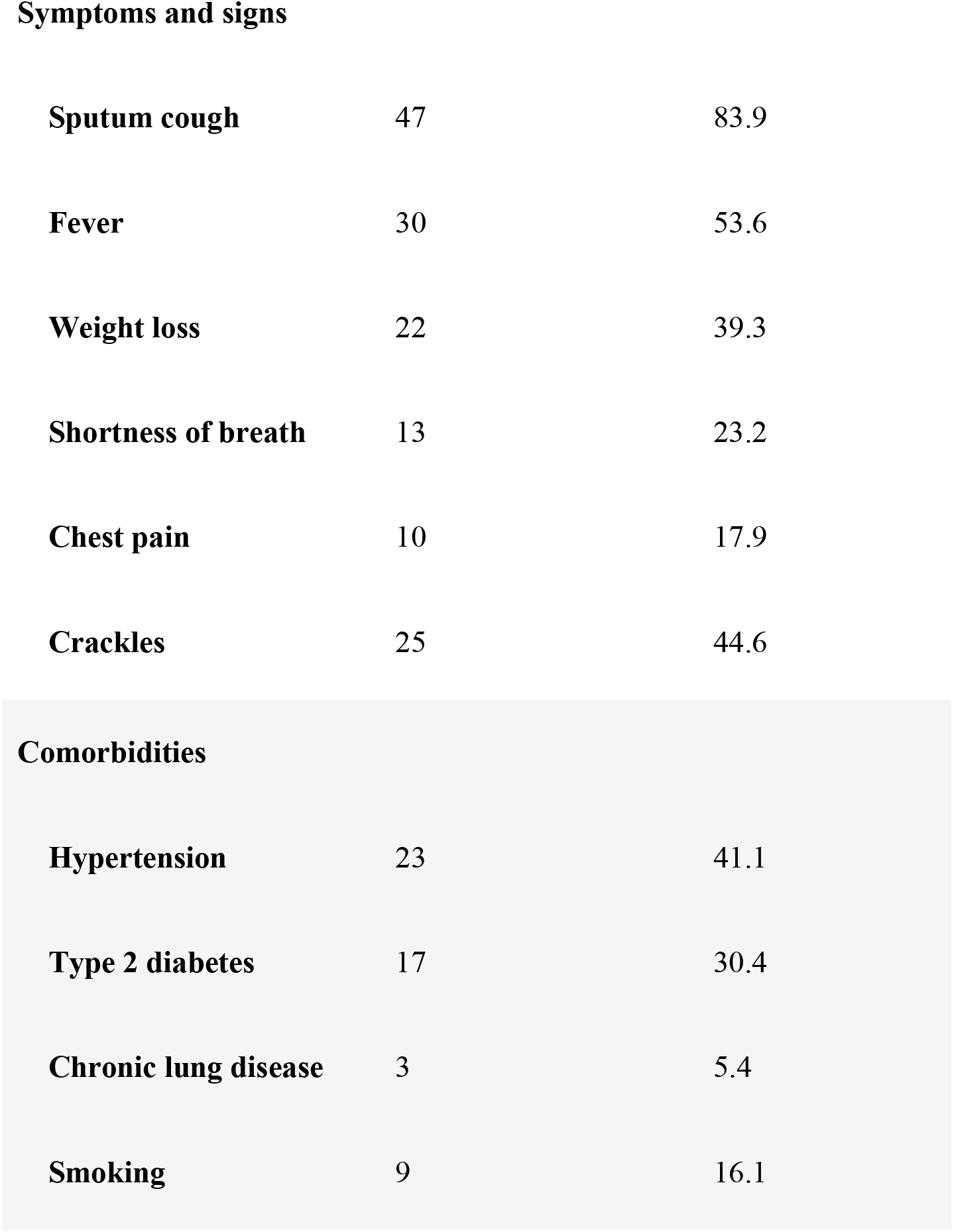
Characteristics of participants with AFB-negative sputum but MGIT-positive bronchoalveolar lavage fluid.

| **Characteristics** | **Frequency (n = 56)** | **Percentage** |
| --- | --- | --- |
| **Sex** |  |  |
| **Male** | 38 | 67.9 |
| **Female** | 18 | 32.1 |
| **Symptoms and signs** |  |  |
| **Sputum cough** | 47 | 83.9 |
| **Fever** | 30 | 53.6 |
| **Weight loss** | 22 | 39.3 |
| **Shortness of breath** | 13 | 23.2 |
| **Chest pain** | 10 | 17.9 |
| **Crackles** | 25 | 44.6 |
| **Comorbidities** |  |  |
| **Hypertension** | 23 | 41.1 |
| **Type 2 diabetes** | 17 | 30.4 |
| **Chronic lung disease** | 3 | 5.4 |
| **Smoking** | 9 | 16.1 |

There were 67.9% of males and 32.1% of females (male/female ratio was 2.1). Common clinical symptoms included cough with sputum (83.9%), followed by fever (53.6%), crackles (44.6%), weight loss (39.3%), shortness of breath (23.2%), and chest pain (17.9%). Frequent comorbidities were hypertension (41.1%), type 2 diabetes (30.4%), and chronic lung disease (5.4%). The smoking rate accounted for 16.1% of participants (Table 1).

There were 48.2% of patients who had injuries in more than one location. The right lung injury rate was more common than the left side (46.4% vs. 28.6%) and the upper lobe injury rate was more than the lower side (51.8% vs. 17.8%). Characteristics of injuries on X-ray included infiltration (64.3%), solidification (21.4%), pleural effusion (10.7%), cavity (8.9%), and nodular (5.4%) (Table 2).

**Table 2.** Chest X-ray characteristics of participants with AFB-negative sputum but MGIT-positive bronchoalveolar lavage fluid.

| **Characteristics** | **Frequency**<br>**(n = 56)** | **Percentage** |
| --- | --- | --- |
| **Location** |  |  |
| **>1 location** | 27 | 48.2 |
| **Right lung** | 26 | 46.4 |
| **Left lung** | 16 | 28.6 |
| **Upper lobe** | 29 | 51.8 |
| **Lower lobe** | 10 | 17.8 |
| **Kind of injury** |  |  |
| **Infiltration** | 36 | 64.3 |
| **Solidification** | 12 | 21.4 |
| **Pleural effusion** | 6 | 10.7 |
| **Cavity** | 5 | 8.9 |
| **Nodular** | 3 | 5.4 |

Gene Xpert MTB/RIF of bronchial lavage fluid had tuberculosis bacteria in 40.8% while MGIT-positive rate was 46.7%. The sensitivity of Gene Xpert MTB/RIF compared to MGIT was 85.7%, and the specificity was 98.4%. The positive predictive value was 97.9% and the negative predictive value was 88.7% (Table 3).

**Table 3.**
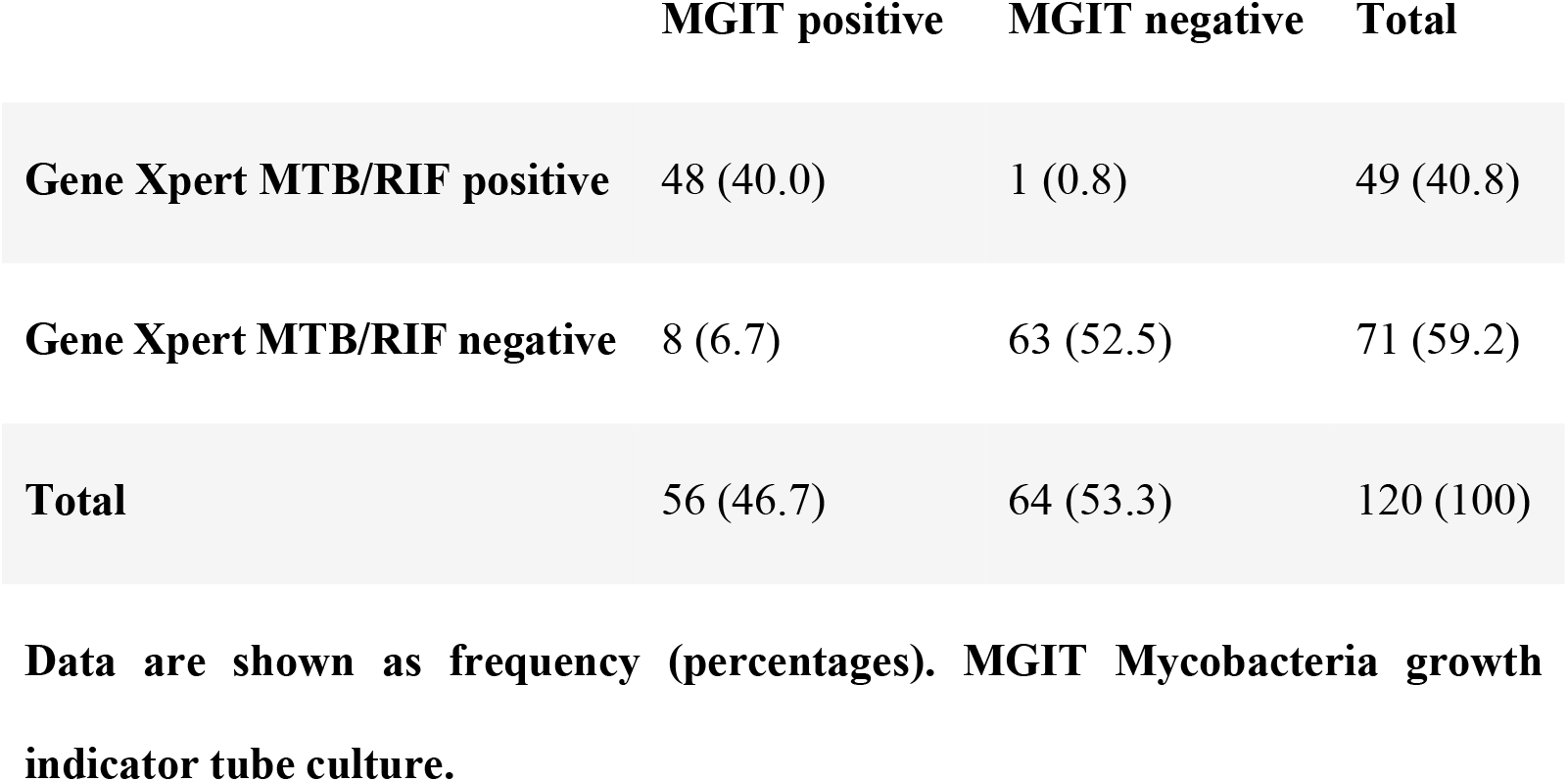
Gene Xpert MTB/RIF compared with MGIT results of bronchial lavage fluid.

|  | MGIT positive | MGIT negative | Total |
| --- | --- | --- | --- |
| **Gene Xpert MTB/RIF positive** | 48 (40.0) | 1 (0.8) | 49 (40.8) |
| **Gene Xpert MTB/RIF negative** | 8 (6.7) | 63 (52.5) | 71 (59.2) |
| **Total** | 56 (46.7) | 64 (53.3) | 120 (100) |

**Data are shown as frequency (percentages). MGIT Mycobacteria growth indicator tube culture.**

Among 49 patients with Gene Xpert MTB/RIF results to detect tuberculosis, the rate of resistance to rifampicin was only 4.1% (Table 4).

**Table 4.** Rifampicin resistance results in Gene Xpert MTB/RIF.

| Characteristics | Frequency<br>(n = 49) | Percentage |
| --- | --- | --- |
| Resistance to rifampicin | 2 | 4.1 |
| Not resistant to rifampicin | 39 | 79.6 |
| Unidentified resistance | 8 | 16.3 |
| Total | 49 | 100 |

## 4. DISCUSSION

The study recruited 120 patients with suspected pulmonary tuberculosis and AFB-negative sputum results. There were 56 patients with MGIT-positive of bronchoalveolar lavage samples whose mean age was 57.8 ± 20.1 and BMI was 21.4 ± 1.8 kg/m2 (Table 1). The male to female ratio in our study was approximately 2.11. This figure was higher than some previous reports for TB in adults, which mentioned ratios of 1.25-1.81 depended on age groups from 15 to over 65, and 1.7 globally independent of age groups ^7,8^. Common comorbidities included hypertension (41.1%), followed by type 2 diabetes (30.4%), and chronic lung disease (5.4%). History of smoking accounted for 16.1%, less than previous studies done in Bangladesh, Pakistan and South Africa, which found rates of current and past smoking from 22.5% up to over 70% ^9,10^.

Common clinical symptoms included cough with sputum (83.9%), followed by fever (53.6%), crackles (44.6%), weight loss (39.3%), shortness of breath (23.2%), and chest pain (17.9%). These findings were in line with literature in the aspect that cough was the most prevalent symptom ^11,12^. Fever rates seemed to vary significantly, with reports of up to 75% and as low as 30% ^11,12^. Haemotypsis was a rare phenomenon in TB patients, though when severe it could become lethal in 5% of cases before effective treatment ^13^. Our study found no patients exhibiting this symptom, which might be due to the low sample size. Previous studies had found the rate of haemotypsis to be 4-8% in TB patients ^11,14^.

In this study, 48.2% of patients had injuries in more than one location on chest X-ray and 64.3% of patients had infiltration (Table 2). This is similar to the findings of Bg et al., which denoted infiltration to be the most common radiological presentation at 50% ^11^. Cavitation was a hallmark of severe disease and increased bacillary burden ^15-17^. In our study, only 8.9% of patients showed this characteristic on chest X ray. Compared to other reports, the rate of cavitation on X ray could be much higher, with different sources estimating the figure to be between 29-87% and 40-87% ^15,18^. Studies analyzing radiological data had given cavitation rates of roughly 30-50% ^11,16^. As this phenomenon was associated with more severe disease and less time to positivity in culture, it might be less prevalent in sputum smear negative patients.

In this study, the sensitivity of Gene Xpert MTB/RIF compared to MGIT was 85.7%, and the specificity was 98.4%. The positive predictive value was 97.9% and the negative predictive value was 88.7%. (Table 3). According to Lee et al., the sensitivity and specificity of Gene Xpert MTB/RIF in bronchial lavage fluid were 81.6% and 100% compared to culture in diagnosing tuberculosis ^19^. Mechal et al. found similar figures at 78.2% and 90.4%, while Elbrolosy et al. reported higher results at 90.2% and 86.9% for sensitivity and specificity in pulmonary TB, respectively ^20,21^. Le Palud’s study showed that bronchoscopy to obtain bronchial lavage fluid or biopsy in patients with suspected tuberculosis showed that Gene Xpert MTB/RIF and culture had nearly equivalent sensitivity (60% versus 66.7%) ^22^. Zhou et al. combined Gene Xpert MTB/RIF with endobronchial ultrasonography with a guide sheath to diagnose TB, achieving increased specificity at 100% but lower sensitivity at 47.26% ^23^.

Among 49 patients with Gene Xpert MTB/RIF results of bronchial lavage fluid detecting tuberculosis bacteria, the rifampicin resistance rate was 4.1%, unknown resistance was 16.3%, and non-resistant accounted for the majority of 79.6% (Table 4). The rate of rifampicin resistance found was within reported range of medical literature. Fadeyi et al. found that rifampicin resistant TB rate was 4.2% in patients with previously treated TB ^24^. Ren et al. screened 53,893 TB patients in Eastern China and found 5.9% cases had rifampicin resistance ^25^. In Ethiopia, Sharew et al. reported a higher rate at 7.1% in 12,981 patients ^26^. The Gene Xpert MTB/RIF method was found to be highly effective in detecting rifampicin resistance, achieving sensitivity and specificity of 88.5-93.3% and 96.4-97.6%, respectively ^27,28^.

## 4. CONCLUSIONS

Pulmonary tuberculosis patients with AFB-negative sputum exhibit a wide range of clinical and radiological characteristics. The Gene Xpert MTB/RIF test on bronchial lavage fluid demonstrates high sensitivity and specificity in diagnosing AFB-negative pulmonary tuberculosis. Additionally, the rate of rifampicin resistance is low.

## Data Availability

All relevant data are within the manuscript and its Supporting Information files.

